# Suicide in young people with Special Educational Needs and Disabilities (SEND) in England 2011 to 2022: A retrospective cohort study

**DOI:** 10.64898/2026.10.30.26366719

**Authors:** Marta Rossa, Isobel Thompson-Ward, Cameron Razieh, Cathryn Rodway, Wendy van Rijswijk, Ania Zylbersztejn, Vahé Nafilyan, Emma Sharland

## Abstract

**Background:** Population-level evidence on suicide mortality among children and young people (CYP) with Special Educational Needs and Disabilities (SEND) is limited. SEND captures a broad and heterogeneous population, including children whose needs are recognised in educational settings regardless of recorded clinical diagnosis.

**Methods:** We analysed 3,959,390 CYP aged 10–18 years in England using GUiE data linked to death registrations. SEND provision was classified as “no support”, “support” or “statement”, using records from academic years 2001/02 to 2010/11. CYP were followed until age 25, death or 31 December 2022. Sex-stratified, age-adjusted Cox models estimated suicide by SEND provision.

**Results:** In the cohort, 1,383,300 CYP (35%) had provision of support and 148,200 (4%) had a statement. During follow-up, 2,620 suicides occurred among 10- to 24-year-olds; 1,190 (45.4%) were among CYP with support and 130 (5%) among those with statements. Compared with males with no support, suicide was higher among males with provision of support (hazard ratio [HR]: 1.46; 95% CI: 1.33–1.61) and statements (HR: 1.32; 95% CI: 1.09–1.61). Among females, support provision was associated with higher risk of suicide (HR: 1.23; 95% CI: 1.05–1.44), while no significant difference was observed for those with statements (HR: 1.15; 95% CI: 0.69–1.92). Suicide rates were highest among CYP with social, emotional and mental health needs.

**Conclusions:** CYP with SEND had elevated risk of suicide, particularly those receiving support without a statutory plan and those with social, emotional and mental health needs, highlighting the need for targeted prevention.

## Introduction

Suicide is a major global public health concern and was the third leading cause of death among 15-29-year-olds worldwide in 2021 (1). In England suicide accounted for 16.4% and 15.3% of all deaths among boys and girls in 2024 respectively (2). Although deaths by suicide in childhood and adolescence are rare in absolute terms, they constitute a substantial proportion of deaths in this age group and have profound consequences for families, schools, communities and services. The Suicide Prevention Strategy for England (2023) identifies children and young people (CYP) as a key group and highlights the importance of understanding suicide among those with complex vulnerabilities, such as neurodevelopmental conditions, mental health difficulties, and those experiencing social exclusion (3). Robust population-level evidence is therefore needed to identify groups at elevated risk of suicide and to inform prevention, service planning and support.

Special Educational Needs and Disabilities (SEND), referred to as special educational needs (SEN) in the historical education records used in this study, refer to children and young people who have difficulties or disabilities which have an impact on their ability to learn (4,5). SEND is a broad administrative and educational classification rather than a clinical diagnosis, encompassing needs related to communication and interaction, cognition and learning, social, emotional and mental health, and sensory or physical difficulties (6). As a result, children with SEND represent a diverse population with a wide range of needs, experiences and support requirements. Children with SEND may receive additional support in school or college, with the level of provision tailored to their individual needs. The number of children recorded as having SEND has increased substantially in England, reaching over 1.7 million children in 2025, with growing concern about variation in access to support and the capacity of services to meet need (7,8).

Previous research suggests that some groups of children and young people whose needs may fall within the SEND system are at increased risk of self-harm, suicidal ideation or suicide-related behaviours. Elevated risks of self-harm and other suicide-related outcomes have been reported among autistic young people and those with attention-deficit hyperactivity disorder (9–12). Social isolation, bullying, mental health difficulties and academic pressures are also recognised as relevant antecedents of self-harm and suicide among young people (13–15). However, existing evidence largely concerns clinical diagnoses, selected service populations, hospital presentations, self-reported outcomes or non-fatal suicidal behaviours, leaving the association between educationally recorded SEND and subsequent suicide mortality unclear. Coroner records and Prevention of Future Death reports have identified autism or autistic traits among some people who died by suicide, including CYP (16), but cannot estimate population-level risk and may be subject to selection, reporting and ascertainment biases. SEND also encompasses broader, more heterogeneous needs than specific clinical diagnoses, including educationally recognised needs without a recorded diagnosis. Given the rarity of suicide among CYP, large linked datasets are needed to estimate mortality across SEND groups. Using population-level linked Census, education and death registration data for England, we examined the association between recorded SEND provision and subsequent suicide mortality, with analyses by sex and type of need.

## Methods

### Data sources

We conducted a population-level retrospective cohort study using the Growing Up in England (GUiE) dataset (17,18), which was linked to Office for National Statistics (ONS) death registrations (19) via a unique census identifier. The GUiE dataset includes individuals residing in England who were linked to the 2011 Census and to education records from the feasibility version of the All Education Dataset for England (AEDE), including records from the National Pupil Database (NPD), Individual Learner Records (ILR) and Higher Education Statistics Agency (HESA) and additional information on vulnerability measures from the Department for Education’s English School Census (ESC) (17,20). In this study, we used information on SEND provision covering academic years 2001/2002 to 2014/2015 for children aged 10 to 25 as of 31 August 2011. The GUIE dataset contains 7,432,115 eligible unique respondents who were in the 2011 Census and could be linked to feasibility AEDE with a 98% linkage rate (17). Ethical approval was obtained from the National Statistician’s Data Ethics Advisory Committee (NSDEC(22(17)).

### Study population

We included CYP aged 10- to 18-years (up to their 19^th^ birthday) on 27 March 2011 (Census Day) who were enrolled in education in England at any time between the academic years 2001/2002 to 2010/2011. An academic year is defined as 1 September to 31 August. Time at risk was defined as the number of days from Census Day to the earliest of their 25^th^ birthday, end of the study period (31 December 2022), or death.

### Exposures

The exposure was the highest level of provision recorded between the 2001/02 and 2010/11 academic years, derived from the Department for Education’s pupil census collections. Provision was categorised as “no support”, “support” or “statement” (Supplementary Table 1). “No support” indicated no recorded SEN/SEND support or statement; “support” indicated support without a statement; and “statement” indicated a recorded statement, representing the highest level of statutory provision (further details are provided in Supplementary Text 1a).

During the study period, provision was recorded under the terminology of special educational needs (SEN), using the definitions and groupings set out in the SEN Code of Practice (2001), before the Children and Families Act 2014 (21,22). We use SEND throughout this paper to align with current terminology. These pre-2014 SEN categories do not map exactly onto the current SEND system; however, the statement category is most closely aligned with children who would now be supported through an EHCP. In our secondary analysis, we investigated SEND type among individuals with a “Statement”, because data on primary SEND type were incomplete for those receiving support only. We disaggregated this category into categories based on previously defined types of SEND from the Department for Education (6,23). Individuals were grouped into one of five SEND types (Supplementary Table 3 and Supplementary Text 1b): (1) cognition and learning difficulties; (2) social, emotional and mental health needs; (3) communication and interaction needs; (4) physical or sensory needs; and (5) unresolved (where multiple SEND types were present in the most recent recorded year or the SEND type was either not recorded or listed as “Other.”).

### Outcome

The primary outcome was death by suicide, defined as death from intentional self-harm for persons aged 10 years and over, and deaths caused by injury or poisoning where the intent was undetermined for those aged 15 years and over (24). We used International Classification of Diseases, Tenth Revision (ICD-10) codes corresponding to intentional self-harm (X60-X84), injury (including, but not limited to, poisoning) of undetermined intent (Y10-Y34) (25).

Our secondary outcome was non-suicide deaths, defined as deaths by all other causes excluding suicide (ICD-10 codes X60-X84 and Y10-Y34).

### Covariates

All sociodemographic variables included in the analysis were sourced from the 2011 Census and were likely to be reported by proxy for the CYP by the main census respondent in the household (26). Covariates included were sex, age, ethnic group, country of birth, main language, National Statistics Socio-Economic Classification (NS-SEC) of the Household Reference Person (HRP), general health status, disability status, unpaid carer status, and type of communal establishment. Categories within all variables can be found in Supplementary Table 4. For all census variables, categories representing missing or inapplicable data (e.g. “Unknown”, “Missing”, or “Not Stated”) were retained as recorded but treated as a single group in analyses (26).

### Statistical analysis

Analyses by SEND provision were stratified by sex given the prior evidence of differences in suicide between sex (14). Analyses by SEND type pooled males and females. Descriptive statistics were reported using mean (standard deviation) for continuous variables and count (percentage) for categorical variables.

Crude and age-standardised mortality rates (ASMRs) for suicide and non-suicide deaths per 100,000 person-years were estimated for our exposure groups (no support, support, statement). Age-standardised mortality rates were estimated by direct standardisation using three-year age bands defined at Census (10-12, 13-15 and 16-18 years), with the age distribution of the study cohort used as the standard population. We calculated 95% confidence intervals (CI) for the ASMRs assuming a Poisson distribution of deaths, using standard errors derived from the observed number of events in each age-sex stratum.

We used cause-specific Cox proportional hazards models to examine suicide risk by SEND status, censoring non-suicide deaths. All models adjusted for age using a natural spline with three knots and boundary knots at the 1st and 99th centiles to capture nonlinearity. Residuals were examined for proportional hazards and model fit. Age-adjusted models were followed by additional models accounting for Census characteristics: ethnic group, country of birth, main language, household NS-SEC, general health, disability, unpaid carer status and communal establishment type. These assessed changes after broader sociodemographic and health adjustment, recognising that some characteristics may lie on, or be influenced by, pathways between SEND provision and suicide. For computational efficiency, we randomly sampled 5% of individuals alive during the study period and retained all who died, applying sampling weights for unequal selection probabilities. Data preparation used Sparklyr 3.3.0 and analysis used R 4.4. Under Department for Education Statistical Disclosure Control requirements, counts were rounded to the nearest 10, percentages to two decimal places, and counts below 10 suppressed (27).

### Secondary analysis

To examine how suicide varied by SEND type, our secondary analysis focused on a sub-cohort of children who had ever received a SEND statement and estimated crude and ASMRs for suicide per 100,000 person-years, implementing a similar methodology to the main results.

For each individual, their SEND type was categorised using the most recently recorded type of need (Supplementary Table 3 and Supplementary Text 1). A secondary analysis defined CYP with autism within the communication and interaction needs group and estimated crude and ASMRs for suicide per 100,000 person-years. This group was of particular interest given the prior literature, and the neurodevelopmental condition being associated with elevated risk of adverse mental health outcomes (28).

### Sensitivity analysis

Given the dataset’s long retrospective coverage, we tested robustness across time periods and cohorts. First, we restricted analyses to academic years 2008/2009– 2010/2011 to assess consistency within a more recent, narrower window. Second, we examined CYP aged 16–18 years on the 2011 Census Day, using SEND records from 2005/2006 onwards to explore identification timing, complexity of needs and age-related changes in SEND profiles. We combined support and statement into “any SEND provision” because few suicide deaths in the restricted cohort prevented stable estimates for separate groups. Third, because many CYP had SEND records across multiple years, we assessed whether using the most frequently rather than most recently recorded SEND type affected its association with suicide.

### Patient and Public Involvement Statement

We established and engaged with two patient advisory groups: group 1 was made up of young adults with experiences of suicidality and/or supporting others with suicidality; group 2 was made up of adults who had lost a child or family member to suicide, or who were supporting children or family members with suicidality either in a personal or professional capacity. We also had a paper review group coordinated by the University of Manchester’s Centre for Mental Health and Safety central patient and public involvement and engagement (PPIE) group.

Members of our patient advisory groups and paper review group were involved in reviewing the results of the study and the reporting of it. All PPIE reviewers also contributed to the dissemination plans for this research.

## Results

### Cohort composition and follow-up

The study population included 3,959,390 individuals aged from 10- to 18-years old and enrolled in education during the academic years 2001/2002 through 2010/2011 (Supplementary Table 1, Supplementary Figure 1). The cohort contributed a total of 46,511,405 person-years at risk. Within our cohort 1,383,300 (35%) were receiving support (814,730 [59%] male) and 148,200 (4%) had a statement (107,920 [73%] male) (Table 1). 1,597,720 (80%) females and 1,612,360 (82%) males were from a White ethnic background (Table 1). Among those with statements (the highest level of provision), a higher percentage of children reported bad health (females: 2,680 [6.7%]; males: 4,590 [4.3%]) compared with those receiving support (females: 4,270 [0.8%]; males: 4,550 [0.6%]) or no support (females: 2,920 [0.2%]; males: 1,510 [0.1%]) (Supplementary Table 4).

**Figure 1.**
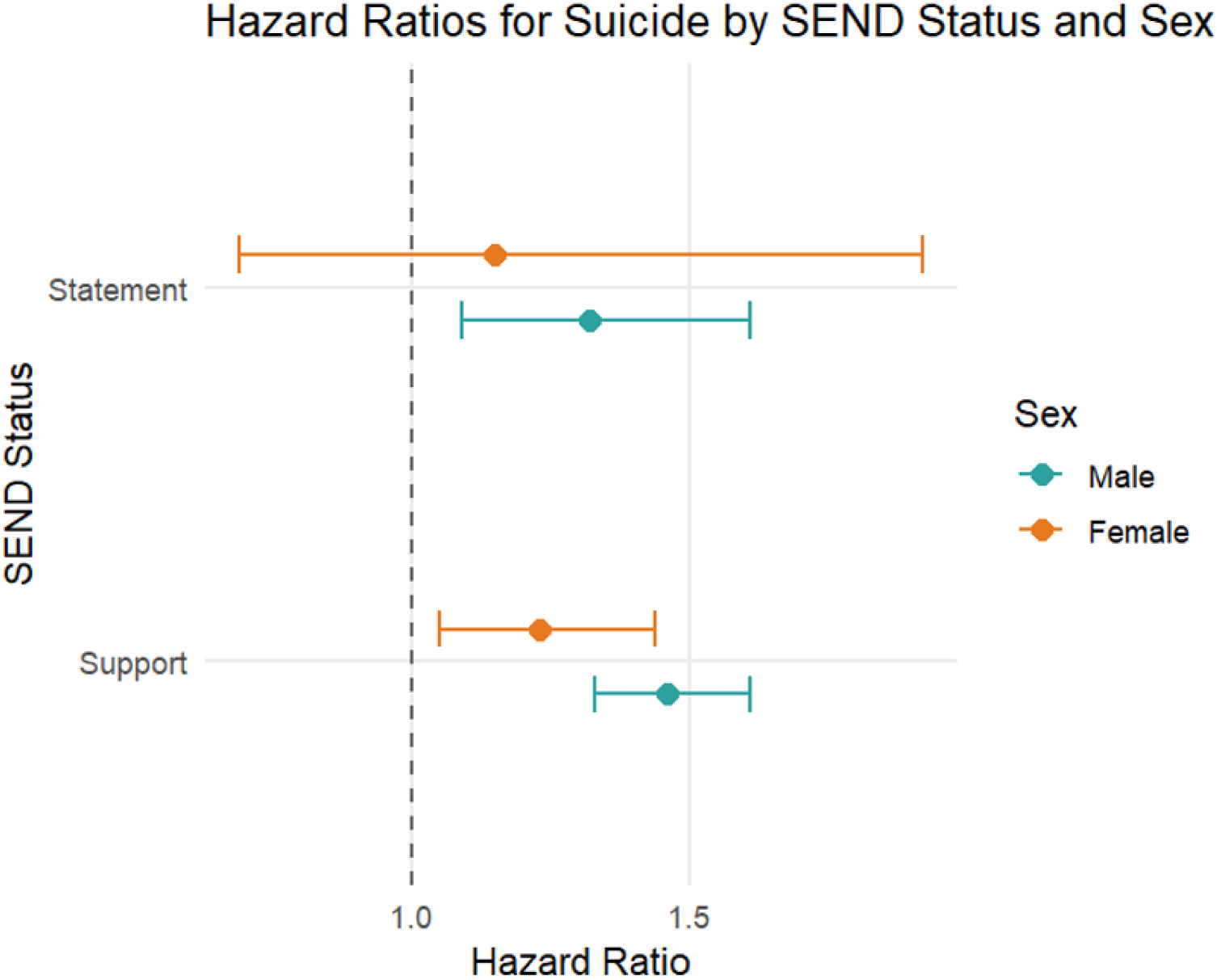
**Hazard ratios (95% CIs) for suicide by sex and SEND status in England, 2011–2022.**

**Table 1:** Demographic characteristics of study population by three levels of SEND.

| Census characteristic | Level | Sex | No support |  |  | Support |  |  | Statement |  |  |
|---|---|---|---|---|---|---|---|---|---|---|---|
|  |  |  | Count (n) | Percentage (%) | Total population (N) | Count (n) | Percentage (%) | Total population (N) | Count (n) | Percentage (%) | Total population (N) |
| Sex | Female |  | 1,361,450 | 56.08 | 2,427,890 | 568,570 | 41.1 | 1,383,300 | 40,280 | 27.18 | 148,200 |
|  | Male |  | 1,066,440 | 43.92 | 2,427,890 | 814,730 | 58.9 | 1,383,300 | 107,920 | 72.82 | 148,20 |
| Ethnic group (aggregated) | Asian/Asian British | Female | 105,230 | 7.73 | 1,361,450 | 44,580 | 7.84 | 568,570 | 2,940 | 7.29 | 40,280 |
|  | Black/African/Caribbean/Black British | Female | 54,890 | 4.03 | 1,361,450 | 33,570 | 5.9 | 568,570 | 1,840 | 4.57 | 40,280 |
|  | Mixed/multiple ethnic groups | Female | 54,680 | 4.02 | 1,361,450 | 24,710 | 4.35 | 568,570 | 1,650 | 4.09 | 40,280 |
|  | Other ethnic group | Female | 11,120 | 0.82 | 1,361,450 | 6,210 | 1.09 | 568,570 | 320 | 0.8 | 40,280 |
|  | White | Female | 1,111,570 | 81.65 | 1,361,450 | 453,460 | 79.75 | 568,570 | 32,690 | 81.16 | 40,280 |
|  | Unknown | Female | 23,960 | 1.76 | 1,361,450 | 6,050 | 1.06 | 568,570 | 840 | 2.09 | 40,280 |
|  | Asian/Asian British | Male | 91,120 | 8.54 | 1,066,440 | 61,850 | 7.59 | 814,730 | 5,380 | 4.98 | 107,920 |
|  | Black/African/Caribbean/Black British | Male | 38,970 | 3.65 | 1,066,440 | 44,410 | 5.45 | 814,730 | 4,870 | 4.51 | 107,920 |
|  | Mixed/multiple ethnic groups | Male | 41,060 | 3.85 | 1,066,440 | 34,800 | 4.27 | 814,730 | 4,580 | 4.25 | 107,920 |
|  | Other ethnic group | Male | 9,700 | 0.91 | 1,066,440 | 8,400 | 1.03 | 814,730 | 700 | 0.65 | 107,920 |
|  | White | Male | 866,060 | 81.21 | 1,066,440 | 656,330 | 80.56 | 814,730 | 89,970 | 83.37 | 107,920 |
|  | Unknown | Male | 19,520 | 1.83 | 1,066,440 | 8,940 | 1.1 | 814,730 | 2,420 | 2.25 | 107,920 |
| Country of birth | Non-UK | Female | 92,390 | 6.79 | 1,361,450 | 44,850 | 7.89 | 568,570 | 1,900 | 4.7 | 40,280 |
|  | UK | Female | 1,245,100 | 91.45 | 1,361,450 | 517,680 | 91.05 | 568,570 | 37,540 | 93.2 | 40,280 |
|  | Missing or not stated | Female | 23,960 | 1.76 | 1,361,450 | 6,050 | 1.06 | 568,570 | 840 | 2.09 | 40,280 |
|  | Non-UK | Male | 80,580 | 7.56 | 1,066,440 | 57,140 | 7.01 | 814,730 | 3,690 | 3.42 | 107,920 |
|  | UK | Male | 966,340 | 90.61 | 1,066,440 | 748,650 | 91.89 | 814,730 | 101,810 | 94.34 | 107,920 |
|  | Missing or not stated | Male | 19,520 | 1.83 | 1,066,440 | 8,940 | 1.1 | 814,730 | 2,420 | 2.25 | 107,920 |

### Suicide deaths and absolute rates

During our follow-up period, a total of 10,640 (0.3%) CYP died; of these, 2,620 (24.6%) died by suicide. This equated to 0.07% of the total cohort. Crude and age-standardised suicide mortality rates by sex and SEND status are presented in Table 2. Among males, estimated crude rates of suicide were higher for those with support (9.98 suicide deaths per 100,000 person-years, 95% CI 9.35, 10.62) and statements (8.95, 95% CI: 7.30, 10.60), compared to those with no support (6.74, 95% CI: 6.29, 7.20). Among females, suicide rates were highest in those with support (3.52, 95% CI: 3.07, 3.97) and lowest in those not requiring support (2.82, 95% CI: 2.56, 3.08). The suicide rate was 3.19 (95% CI: 1.79, 5.26) in those with statements (Supplementary Table 7).

**Table 2.** Counts and percentage of children with Special Educational Needs in study population & crude and age-standardised rates per 100,000 person-years for death by suicide.

| **Special Educational Needs category** | **Sex** | **Count (n)** | **Percentage (%)** | **Total population (n)** | **Count of suicides (n)** | **Crude rate per 100,000 person-years [95% CI]** | **Age Standardised Mortality Rate (ASMR) per 100,000 person-years [95% CI]** |
|---|---|---|---|---|---|---|---|
| No Support | Female | 1,361,450 | 56.08 | 2,427,890 | 450 | 2.82 [2.56-3.08] | 2.82 [2.56-3.08] |
| Support | Female | 568,570 | 41.1 | 1,383,300 | 240 | 3.52 [3.07-3.97] | 3.51 [3.06-3.96] |
| Statement | Female | 40,280 | 27.18 | 148,200 | 20 | 3.19 [1.79-5.26] | 3.18 [1.78-5.25] |
| No Support | Male | 1,066,440 | 43.92 | 2,427,890 | 850 | 6.74 [6.29-7.2] | 6.76 [6.31-7.22] |
| Support | Male | 814,730 | 58.9 | 1,383,300 | 960 | 9.98 [9.35-10.62] | 9.95 [9.32-10.58] |
| Statement | Male | 107,920 | 72.82 | 148,200 | 110 | 8.95 [7.3-10.6] | 8.77 [7.15-10.39] |

### Age-adjusted hazard ratios

In Cox proportional hazards models adjusted for age, males receiving support had a 46% higher risk of suicide than males with no support (HR 1.46, 95% CI 1.33 to 1.61). Males with statements had a 32% higher risk of suicide than males with no support (HR 1.32, 95% CI 1.09 to 1.61). Among females, receipt of support was associated with a 23% higher risk of suicide compared with no support (HR 1.23, 95% CI 1.05 to 1.44). Among females, there was no clear evidence of a difference in suicide risk between those with statements and those with no support (HR 1.15, 95% CI 0.69 to 1.92). Findings were largely unchanged after additional adjustment for ethnicity, main language, communal establishment and unpaid carer status (Supplementary Table 10). Greater attenuation was observed following adjustment for health, disability status and household NS-SEC, particularly among individuals with statements, whereas associations among individuals receiving support remained largely unchanged.

### Secondary analysis; analysis of SEND type

Among CYP with statements (n= 148,200), 39% (n= 58,120) had a primary type of cognition and learning difficulties, 28% (n= 42,040) with communication and interaction (60% (n= 25,270) of those had autism), 16% (n= 23,690) with social, emotional and mental health needs, and 10% (n= 14,870) with sensory and/or physical needs. An additional 6% (n= 9,490) of children were classified as having ‘Unresolved’ SEND types (see Table 3). Disability status varied by SEND type. Activity limitation was most common among CYP with sensory and/or physical needs (11,630; 78.23%) and communication and interaction needs (22,870; 54.4%), whereas most pupils with social, emotional and mental health needs (14,700; 62.04%) and unresolved SEND type (5,450; 57.48%) reported no activity limitation. For broader SEND-type characteristics see Supplementary Table 5; and autism-specific characteristics see Supplementary Table 6.

**Table 3.** Counts and percentage among CYP with SEND statement by SEND types, and crude and age-standardised rates per 100,000 person-years for death by suicide.

| Type of Special Educational Need and Disability | Count (n) | Percentage (%) | Total population (n) | Count of suicides (n) | Crude rate per 100,000 person-years [95% CI] | ASMR per 100,000 person-years [95% CI] |
|---|---|---|---|---|---|---|
| Communication and interaction | 42040 | 28.36 | 148200 | 30 | 6.48 [4.24-8.73] | 6.63 [4.52-9.37] |
| Cognition and learning | 58120 | 39.22 | 148200 | 40 | 5.31 [3.58-7.05] | 5.18 [3.59-7.21] |
| Social, emotional and mental health | 23690 | 15.98 | 148200 | 40 | 15.5 [10.87-20.13] | 14.67 [10.57-19.81] |
| Sensory and/or physical needs | 14870 | 10.03 | 148200 | n<10<sup>1</sup> | - | - |
| Unresolved<sup>2</sup> | 9490 | 6.4 | 148200 | 10 | 11.7 [5.34-18.05] | 12.84 [6.73-22.13] |

***Note:*** *<sup>1</sup>Suppressed due to small counts (n < 10). <sup>2</sup>Unresolved (where multiple SEND types were present in the most recent recorded year or the SEND type was either not recorded or listed as "Other.")*

Of the 2,620 deaths attributed to suicide in CYP, 130 (5%) of these were among children with statements. The estimated crude suicide rates (Table 3) were highest among those with social, emotional, and mental health needs (15.5 per 100,000 person-years; 95% CI: 10.87, 20.13), followed by those with communication and interaction (6.48, 95% CI: 4.24, 8.73) and cognition and learning difficulties (5.31, 95% CI: 3.58, 7.05). For context, the crude suicide rate among CYP with no support was 4.55 per 100,000 person-years (95% CI: 4.30 to 4.79). Within the communication and interaction group, CYP with autism had a crude suicide rate of 7.13 per 100,000 person-years (95% CI: 4.28 to 9.98) (Supplementary Table 9).

### Sensitivity analyses

Findings were broadly consistent across sensitivity analyses (Supplementary Tables 11 and 12; Supplementary Texts 2 and 3). Restricting exposure to 2008/09–2010/11 modestly attenuated the association among males with statements (HR 1.20, 95% CI 0.98–1.48), while estimates for support remained similar among males (HR 1.40, 95% CI 1.27–1.55) and were stronger among females (HR 1.40, 95% CI 1.18–1.66). Among those aged 16–18 years, combining support and statements because of small counts showed elevated suicide risk in males (HR 1.44, 95% CI 1.23–2.01) and females (HR 1.48, 95% CI 1.09–2.01). Ranking of suicide rates by SEND type was unchanged using the most frequently recorded type (Supplementary Table 13; Supplementary Text 4).

## Discussion

This study provides comprehensive population-level analysis of suicide among CYP with SEND in England. Our findings indicate that males who received any SEND provision (support or statement) and females with support only had significantly higher risk of dying by suicide compared to their peers who did not have a record of SEND provision. This work provides key insights into high-risk groups of CYP in England and underscores the need for targeted prevention strategies.

Previous research corroborates our findings, highlighting the vulnerability of CYP with SEND. Self-harm and mental illness are commonly reported antecedents of suicide among young people (15), and studies using smaller cohorts have shown higher rates of self-harm and suicidal ideation among those with SEND (9,29). This may reflect overlapping vulnerabilities within the SEND population: children with learning disabilities, autism and ADHD are more likely to experience social isolation and bullying, which are associated with anxiety, depression and poorer wellbeing (12,29). These risks may be further compounded by other challenges like housing instability, financial hardship, and limited social support which are also well-established predictors linked with psychological distress, loneliness, and reduced wellbeing among adolescents (13,30).

Notably, the highest risk of suicide appears among those receiving support rather than those with a formal statement. In our cohort, approximately 35% of children had received support at any given time, compared to only 4% with a formal statement, indicating that only a very small proportion were accessing higher-level, coordinated support. These proportions describe recorded provision and do not establish whether support met individual needs.

Analysis by the Health Foundation indicates that children with SEND are disproportionately represented in lower-income households (31). This socioeconomic context is relevant when considering the needs of CYP across levels of SEND provision. Such inequity is concerning given the elevated risk of poor mental health and suicide among children from low-income backgrounds. According to current statutory guidance, EHCPs are intended for children whose needs exceed what can be met through the SEND support provided by the school (5). Yet in practice, the boundary between these groups is often blurred. Many children receiving SEND support may have complex needs similar to those with EHCPs, but lack access to the same level of coordinated services (8). This gap and inequalities in provision may contribute to their increased vulnerability.

In sex-stratified analyses, support and statements were associated with higher suicide risk among males compared with those with no recorded SEND provision. Among females, support only was associated with higher risk, while the estimate for statements provided no clear evidence of a difference from those with no recorded provision. These findings do not establish that the associations differed between males and females. The small number of suicide deaths among females with statements limits the precision of the estimate, and findings for this group should therefore be interpreted cautiously. Related evidence from a Swedish population-based study found that suicide incidence following medically treated self-harm was higher among autistic males than autistic females (32). However, differences in the population and exposure definitions limit direct comparison with our findings.

Statements capture a broad range of needs, including physical, sensory, cognitive, social, emotional and neurodevelopmental difficulties. For some less visible neurodevelopmental needs, diagnostic or identification bias may play a role: previous research suggests that autistic girls may be under-identified because their difficulties can be less visible or masked through compensatory behaviours (33). This may mean that some girls with less visible needs are less likely to receive formalised support, although this explanation is unlikely to apply equally across all statement types. Girls are also more likely to seek help for emotional distress and to talk openly about mental health, which can lead to earlier support (15,34), whereas boys with SEND, especially those with communication or behavioural challenges, may face greater social isolation, increasing their vulnerability. Our findings do highlight however, the need to strengthen support at both the support and statement level, particularly for boys. Despite receiving formal support, males with statements still showed an elevated risk of suicide, suggesting that the higher-level provisions may not fully meet their needs.

While our study highlights increased risk among CYP with SEND, this risk is not limited to one type of SEND but extends across a broad spectrum of need. This is particularly important given the heterogeneity of the SEND population, which includes CYP with diverse cognitive, emotional, and physical needs. Our secondary analysis revealed that suicide rates varied by SEND type among CYP with statements, with the highest estimated rate observed among those with social, emotional and mental health (SEMH) needs. The Department for Education reported that boys with SEMH (but no statement) were 3.8 times more likely to be actively excluded than a children without SEND, and girls with SEMH were around 3.0 times more likely after controlling for other factors (35). This suggests that children with SEMH are particularly vulnerable, especially due to challenges with emotional regulation (36).

### Strengths and Limitations

Key strengths include population coverage of children and young people in England through linked administrative data and follow-up into early adulthood, enabling robust estimation of suicide rates across SEND provision groups and extending evidence from smaller cohorts. Historical pre-2014 SEN records provided a broadly consistent statutory framework before the transition from statements to EHCPs, supporting exposure-group comparability and longer-term follow-up. A comparison group without identified mental or physical difficulties or disabilities affecting education provided a reference for SEND-associated risk. Examining provision levels rather than SEND status alone captured differences in support received, clarifying how needs and provision relate to suicide risk.

Several limitations warrant consideration. First, the data predate the Children and Families Act 2014 reforms, limiting comparability with current SEND provision. Statements and EHCPs both provide statutory support for complex needs, but EHCPs were designed to integrate education, health and social care and extend support to age 25. However, evidence suggests similar implementation challenges persist, with provision remaining largely education focused (37).

Second, incomplete SEND-type recording restricted analyses to children with statements, while few suicide deaths within categories limited precision and sex-specific and other subgroup analyses.

A third limitation concerns the dynamic nature of both SEND status and the type of identified need. Both SEND status and the type of identified need, such as autism, ADHD, or emotional and behavioural difficulties, can change over time, particularly in younger children, as their developmental profiles evolve. Although sensitivity analyses were conducted to address this, some residual bias may persist. Children’s primary SEND needs often vary across school years, and many have co-occurring difficulties, reflecting both developmental changes and how schools assess, record and respond to those needs.

### Implications and Future directions

The findings revealed that CYP receiving SEND provision but without a formal statutory plan may face an elevated risk of suicide. Policy and research efforts must also recognise the intersectional vulnerabilities faced by this population. While autism and ADHD are receiving increasing attention, broader SEND categories remain under-researched, constraining understanding of their long-term impacts. Future studies should explore how co-occurring needs may compound risk pathways, as well as how these dynamics differ by sex and are shaped by protective factors.

## Conclusion

Children and young people with SEND face a significantly elevated risk of suicide, particularly those with emotional and behavioural difficulties. These findings highlight the need for inclusive, tailored mental health support and suicide prevention strategies that address the unique challenges faced by this vulnerable population.

## Acknowledgements

We would like to thank the participants of our two public and patient advisory groups. Our groups comprised of young adults with experiences of suicidality or supporting others with suicidality, and secondly adults who had lost a child or family member to suicide, or who were supporting children or family members with suicidality either in a personal or professional capacity. Members of our public and patient advisory groups reviewed the results of the analysis and provided feedback. We would like to thank Papyrus UK and The Mix for facilitating these workshops.

We would also like to thank members of the Mutual Support for Mental Health Research (MS4MH-R) Patient and Public Involvement and Engagement group affiliated with the NIHR Greater Manchester Patient Safety Research Collaboration and Centre for Mental Health and Safety at The University of Manchester for their contribution to the interpretation of results and reviewing of publications.

Cathryn Rodway is supported by the National Institute for Health Research (NIHR) Applied Research Collaboration – East Midlands (ARC-EM), the NIHR Leicester Biomedical Research Centre (BRC), and Leicester British Heart Foundation Centre of Research Excellence.

## Author contributions

Emma Sharland and Isobel Ward conceived, acquired funding and designed the study. Marta Rossa led the data analysis with support from: Isobel Ward, Emma Sharland and Cameron Razieh. Marta Rossa, Emma Sharland, Isobel Ward, Cathryn Rodway, Wendy van Rijswijk, and Vahé Nafilyan quality assured the outputs and the interpretation of them. Marta Rossa led the write-up of the draft manuscript. Emma Sharland, Isobel Ward, Cameron Razieh, Cathryn Rodway and Vahé Nafilyan contributed to critical revision of the manuscript. All authors approved the final manuscript. Microsoft Copilot was used to improve the grammar, language and flow of the manuscript.

## Funding

This project is funded by the <u>National Institute for Health and Care Research (NIHR)</u> <u>Policy Research Programme (NIHR205990)</u>. The views expressed are those of the author(s) and not necessarily those of the NIHR or the Department of Health and Social Care.

## Ethics approval and consent to participate

Ethical approval was obtained from the National Statistician’s Data Ethics Advisory Committee (NSDEC(22(17))). Informed consent was not required for this study.

## Competing interests/conflict of interest

The authors do not report any conflicts of interest.

## Data availability statement

The source data are not publicly available and are subject to controlled access due to their sensitive nature. Census 2011 and death registration data are available through the Secure Research Service (SRS). Details of the application requirements and process, and the use of data, are available at https://www.ons.gov.uk/aboutus/whatwedo/statistics/requestingstatistics/secureresearchservice

## Code availability statement

Code supporting the analysis may be made available following the relevant ONS approval processes.

## Notes

### Competing Interest Statement

The authors have declared no competing interest.

### Author Declarations

The National Statisticians Data Ethics Advisory Committee gave ethical approval for this work NSDEC 22 17.

## References

1. The World Health Organisation. Suicide worldwide in 2021 global health estimates. 2025; Available from: https://www.who.int/publications/b/77047

2. Office for National Statistics. Deaths registered in Englan and Wales: 2023. 2024; Available from: https://www.ons.gov.uk/peoplepopulationandcommunity/birthsdeathsandmarriages/deaths/bulletins/deathsregistrationsummarytables/2023#leading-causes-of-death

3. Department of Health and Social Care. Suicide prevention strategy for England: 2023 to 2028 [Internet]. 2023. ([cited 2024 Jun 27]). Available from: https://www.gov.uk/government/publications/suicide-prevention-strategy-for-england-2023-to-2028

4. Department for Education and Edward Timpson CBE KC. Reforms for children with SEN and disabilities come into effect. [internet] [Internet]. 2014; Available from: https://www.gov.uk/government/news/reforms-for-children-with-sen-and-disabilities-come-into-effect

5. UK Government. Children and Families Act 2014: Chapter 6 [Internet]. (London: legislation.gov.uk). Available from: https://www.legislation.gov.uk/ukpga/2014/6/contents

6. Department for Education. The four broad areas of need [Internet]. Help for early years providers; 2024 [Internet]. Available from: https://help-for-early-years-providers.education.gov.uk/support-for-practitioners/send-assessment/four-broad-areas

7. Department for Education. Special educational needs in England: Academic year 2024/25 [Internet]. 2025. Available from: https://explore-education-statistics.service.gov.uk/find-statistics/special-educational-needs-in-england/2024-25

8. Committee E. Solving the SEND Crisis. 2024;

9. O’Grady SM, Hinshaw SP. Long-term outcomes of females with attention-deficit hyperactivity disorder: Increased risk for self-harm. Br J Psychiatry. 2021;218(1):64–5.

10. Widnall E, Epstein S, Polling C, Velupillai S, Jewell A, Dutta R, et al. Autism spectrum disorders as a risk factor for adolescent self-harm : a retrospective cohort study of 113, 286 young people in the UK. BMC Med [Internet]. 2022;1–14. Available from: 10.1186/s12916-022-02329-w

11. Mars B, Heron J, Klonsky ED, Moran P, Connor RCO, Tilling K, et al. Predictors of future suicide attempt among adolescents with suicidal thoughts or non-suicidal self-harm : a population-based birth cohort study. The Lancet Psychiatry [Internet]. 6(4):327–37. Available from: 10.1016/S2215-0366(19)30030-6

12. Kwan C, Gitimoghaddam M, Collet JP. Effects of Social Isolation and Loneliness in Children with Neurodevelopmental Disabilities: A Scoping Review. Brain Sci. 2020 Oct;10(11).

13. Filia K, Teo SM, Brennan N, Freeburn T, Baker D, Browne V, et al. Interrelationships between social exclusion, mental health and wellbeing in adolescents: Insights from a national Youth Survey. Epidemiol Psychiatr Sci. 2025;34.

14. Hughes JL, Horowitz LM, Ackerman JP, Adrian MC, Campo J V., Bridge JA. Suicide in young people: screening, risk assessment, and intervention. Bmj. 2023;1–16.

15. Rodway C, Tham SG, Ibrahim S, Turnbull P, Kapur N, Appleby L. Children and young people who die by suicide: childhood-related antecedents, gender differences and service contact. BJPsych Open. 2020;6(3):1–9.

16. Sharland E, Wallace E, Revie L, Ward I, Rodway C, Ayoubkhani D. A thematic analysis of Prevention of Future Death reports for children who died by suicide in England and Wales : January 2015 to November 2023. 2025;1–6.

17. Office for National Statistics. Growing Up in England (GUiE). Available from: https://www.ons.gov.uk/peoplepopulationandcommunity/educationandchildcare/methodologies/growingupinenglandguie#wave-1-linkage-methodology-and-wave-2-join

18. ADRUK. Growing Up in England. Available from: https://www.adruk.org/data-access/flagship-datasets/growing-up-in-england/

19. Office for National Statistics (ONS). User guide to mortality statistics [Internet]. Available from: https://www.ons.gov.uk/peoplepopulationandcommunity/birthsdeathsandmarriages/deaths/methodologies/userguidetomortalitystatisticsjuly2017

20. Creation PS, Process L, Quality L. Growing Up in England (GUiE) Wave 1 and Wave 2 Data Quality Manual. 2011;

21. Department for Education. Special educational needs (SEN) code of practice [Internet]. Available from: https://www.gov.uk/government/publications/special-educational-needs-sen-code-of-practice

22. UK Government. Children with special educational needs and disabilities (SEND) [Internet]. Available from: https://www.gov.uk/children-with-special-educational-needs/support-before-september-2014

23. UK Government. SEND code of practice: 0 to 25 years. Available from: https://www.gov.uk/government/publications/send-code-of-practice-0-to-25

24. Office for National Statistics. Suicides in the UK: 2014 registrations [Internet]. 2016. Available from: https://www.ons.gov.uk/peoplepopulationandcommunity/birthsdeathsandmarriages/deaths/bulletins/suicidesintheunitedkingdom/2014registrations#methodological-changes

25. World Health Organisation. International Classification of Diseases (ICD) [Internet]. Available from: https://www.who.int/standards/classifications/classification-of-diseases

26. Office for National Statistics (ONS). Quality and methods [Internet]. Available from: https://www.ons.gov.uk/census/2011census/2011censusdata/2011censususer guide/qualityandmethods

27. Statistics N. Statistical Disclosure Control Policy for DfE data. 2026;(April).

28. O’Halloran L, Coey P, Wilson C. Suicidality in autistic youth: A systematic review and meta-analysis. Clin Psychol Rev [Internet]. 2022;93:102144. Available from: https://www.sciencedirect.com/science/article/pii/S0272735822000290

29. Accardo AL, Neely LC, Pontes NMH, Pontes MCF. Bullying Victimization is Associated with Heightened Rates of Anxiety and Depression Among Autistic and ADHD Youth: National Survey of Children’s Health 2016-2020. J Autism Dev Disord. 2025 Oct;55(10):3605–21.

30. Blackburn CM, Spencer NJ, Read JM. Prevalence of childhood disability and the characteristics and circumstances of disabled children in the UK: secondary analysis of the Family Resources Survey. BMC Pediatr. 2010 Apr;10:21.

31. The Health Foundation. Inequalities in support for special educational needs by free school meal eligibility [Internet]. 2025. Available from: https://www.health.org.uk/evidence-hub/education/schools/inequalities-in-support-for-special-educational-needs-by-free-school

32. Hull L, Stark I, Ohlis A. Sex differences in self-harm and suicide in young autistic adults. 2024;(April):223–33.

33. Lai MC, Lombardo M V., Auyeung B, Chakrabarti B, Baron-Cohen S. Sex/Gender Differences and Autism: Setting the Scene for Future Research. J Am Acad Child Adolesc Psychiatry [Internet]. 2015;54(1):11–24. Available from: 10.1016/j.jaac.2014.10.003

34. Rickwood D, Deane FP, Wilson CJ, Ciarrochi J. Young people’s help-seeking for mental health problems. Aust e-Journal Adv Ment Heal. 2005;4(3):218–51.

35. Department for Education. Edward Timpson publishes landmark exclusions review [Internet]. 2019. Available from: https://www.gov.uk/government/news/edward-timpson-publishes-landmark-exclusions-review

36. Colmenero-Navarrete L, García-Sancho E, José M. S. Relationship Between Emotion Regulation and Suicide Ideation and Attempt in Adults and Adolescents: A Systematic Review. Arch Suicide Res. 2021 Nov 25;26:1–34.

37. Ahad A, Thompson AM, Hall KE. Identifying service users’ experience of the education, health and care plan process: A systematic literature review. Rev Educ. 2022;10(1).

