## Supplementary Tables and Figures for "Suicide in young people with Special Educational Needs and Disabilities (SEND) in England 2011 to 2022: A retrospective cohort study"

**Supplementary Material**

### 1. Supplementary Tables

#### **Supplementary Table 1**: Main analysis: Special Educational Needs and Disabilities: Status of SEND and its categories

| **SEND provision aggregated** | **Description** | **SEND categories pre 2014 Children and Families Act reform** |
| --- | --- | --- |
| No support | A child has no record of any type of SEN provision | No SEN support |
| Support | Indicates that a child has a record of SEN provision but never received a statement | School Action, School Action Plus (SAP), SAP and Statutory Assessment, SAP or Early Years Intervention |
| Statement | A child has a record of the highest level of SEND provision | Statement of SEN |

#### **Supplementary Table 2**: Sample flow

| **Sample** | **Count, individuals** |
| --- | --- |
| All people enumerated in Census 2011 | 56,963,120 |
| Usual resident in Census 2011 | 56,075,910 |
| Living in England in Census 2011 | 53,012,460 |
| Number of unique Administrative Data Research (ADR) IDs in Growing Up in England (GUiE) dataset in wave 3.1 (includes data from academic years 2010/2011 to 2014/2015) | 10,540,550 |
| Number of unique ADR IDs in GUiE dataset in wave 3.2 (includes data from academic years 2005/2006 to 2009/2010) | 10,245,150 |
| Number of unique ADR IDs in GUiE dataset in wave 3.2 Pupil Level Annual School Census (PLASC) (includes data from academic years 2001/2002 to 2005/2006) | 10,330,090 |
| Number of unique ADR IDs with a link to Census 2011 in GUiE wave 3.1 | 3,479,460 |
| Number of unique ADR IDs with a link to Census 2011 in GUiE wave 3.2 | 5,520,590 |
| Number of unique ADR IDs with a link to Census 2011 in GUiE wave 3.2_plasc | 6,482,790 |
| Number of unique ADR IDs with a link to Census 2011 in combined waves (3.1, 3.2 and 3.2_plasc) | 6,745,390 |
| Number of unique ADR IDs with a link to Census 2011 found in GUiE dataset where Census 2011 information are linked to educational dataset for academic years from 2001/2002 to 2014/2015 | 7,432,530 |
| Number of unique ADR IDs with a link to Census 2011 found in GUiE dataset where Census 2011 information are linked to educational dataset for academic years from 2001/2002 to 2010/2011 | 6,720,070 |
| Number of unique ADR IDs linked to multiple Census 2011 IDs found in a sample (of 6,720,070 unique ADR IDs linked to Census in academic years 2001/2002 to 2010-2011). These were removed from the sample. | 200 |
| Main analysis final sample: number of unique ADR IDs linked to Census 2011 and combined three waves; restricted to academic years form 2001/ 2002 to 2010/2011; and restricted to age 10 to 18 (inclusive) on the 2011 Census day. | 3,959,390 |
| Sensitivity analysis final sample: number of unique ADR IDs linked to Census 2011 and combined three waves; restricted to academic years from 2008/2009 to 2010/2011; and restricted to age 10 to 18 (inclusive) on the 2011 Census day. | 3,751,580 |

#### **Supplementary Table 3**: Secondary analysis: Lookup table - Special Educational Needs and Disability types and its categories

| **SEND Types** | **Description** | **Code** |
| --- | --- | --- |
| Communication and interaction | Speech, language & communication needs | SLCN |
| Communication and interaction | Speech, language & communication needs | SLCD |
| Communication and interaction | Autistic spectrum disorder | ASD |
| Communication and interaction | Autistic spectrum disorder | AUT |
| Cognition and learning | Moderate learning difficulty | MLD |
| Cognition and learning | Severe learning difficulty | SLD |
| Cognition and learning | Profound & multiple learning difficulty | PMLD |
| Cognition and learning | Specific learning difficulty | SPLD |
| Cognition and learning | Learning difficulty | LD |
| Social, emotional and mental health difficulties | Social, emotional & mental health | SEMH |
| Sensory and/or physical needs | Visual impairment | VI |
| Sensory and/or physical needs | Hearing impairment | HI |
| Sensory and/or physical needs | Multisensory impairment | MSI |
| Sensory and/or physical needs | Physical disability | PD |
| Unresolved | Other | OTH |
| Unresolved | A clear primary SEN type could not be determined using the approach; or if multiple SEN types occurred in the most recent year and their historical records showed an equal frequency of different types; or if categorised SEN type was either not recorded |  |

#### **Supplementary Table 4:** Main analysis: Sociodemographic characteristics of study population by three levels of SEND

| **Census characteristic** | **Level** | **Sex** | **No support** | | | **Support** | | | **Statement** | | |
| --- | --- | --- | --- | --- | --- | --- | --- | --- | --- | --- | --- |
|  |  |  | **Count (n)** | **Percentage (%)** | **Total population (N)** | **Count (n)** | **Percentage (%)** | **Total population (N)** | **Count (n)** | **Percentage (%)** | **Total population (N)** |
| Disability status | Day to day activities limited a lot/a little | Female | 25,980 | 1.91 | 1,361,450 | 30,920 | 5.44 | 568,570 | 22,450 | 55.73 | 40,280 |
|  | Day to day activities not limited | Female | 1,311,510 | 96.33 | 1,361,450 | 531,600 | 93.5 | 568,570 | 16,990 | 42.17 | 40,280 |
|  | Unknown | Female | 23,960 | 1.76 | 1,361,450 | 6,050 | 1.06 | 568,570 | 840 | 2.09 | 40,280 |
|  | Day to day activities limited a lot/a little | Male | 17,540 | 1.64 | 1,066,440 | 46,550 | 5.71 | 814,730 | 52,640 | 48.78 | 107,920 |
|  | Day to day activities not limited | Male | 1,029,380 | 96.52 | 1,066,440 | 759,230 | 93.19 | 814,730 | 52,860 | 48.98 | 107,920 |
|  | Unknown | Male | 19,520 | 1.83 | 1,066,440 | 8,940 | 1.1 | 814,730 | 2,420 | 2.25 | 107,920 |
| Health status | Very bad health | Female | 570 | 0.04 | 1,361,450 | 980 | 0.17 | 568,570 | 1,470 | 3.65 | 40,280 |
|  | Bad health | Female | 2,920 | 0.21 | 1,361,450 | 4,270 | 0.75 | 568,570 | 2,680 | 6.66 | 40,280 |
|  | Fair health | Female | 21,430 | 1.57 | 1,361,450 | 21,720 | 3.82 | 568,570 | 7,640 | 18.96 | 40,280 |
|  | Good health | Female | 259,730 | 19.08 | 1,361,450 | 148,940 | 26.2 | 568,570 | 12,880 | 31.97 | 40,280 |
|  | Very good health | Female | 1,052,840 | 77.33 | 1,361,450 | 386,620 | 68 | 568,570 | 14,770 | 36.66 | 40,280 |
|  | Unknown | Female | 23,960 | 1.76 | 1,361,450 | 6,050 | 1.06 | 568,570 | 840 | 2.09 | 40,280 |
|  | Very bad health | Male | 390 | 0.04 | 1,066,440 | 1,050 | 0.13 | 814,730 | 2,200 | 2.04 | 107,920 |
|  | Bad health | Male | 1,510 | 0.14 | 1,066,440 | 4,550 | 0.56 | 814,730 | 4,590 | 4.25 | 107,920 |
|  | Fair health | Male | 12,140 | 1.14 | 1,066,440 | 24,470 | 3 | 814,730 | 15,380 | 14.25 | 107,920 |
|  | Good health | Male | 175,320 | 16.44 | 1,066,440 | 187,580 | 23.02 | 814,730 | 34,750 | 32.2 | 107,920 |
|  | Very good health | Male | 857,550 | 80.41 | 1,066,440 | 588,130 | 72.19 | 814,730 | 48,580 | 45.02 | 107,920 |
|  | Unknown | Male | 19,520 | 1.83 | 1,066,440 | 8,940 | 1.1 | 814,730 | 2,420 | 2.25 | 107,920 |
| Household language | All adults speak English as a main language | Female | 1,211,040 | 88.95 | 1,361,450 | 490,200 | 86.22 | 568,570 | 35,690 | 88.61 | 40,280 |
|  | At least one (but not all) adults speak/s English as a main language | Female | 62,750 | 4.61 | 1,361,450 | 30,070 | 5.29 | 568,570 | 2,040 | 5.07 | 40,280 |
|  | No adults speak English as a main language but at least one child does | Female | 42,520 | 3.12 | 1,361,450 | 23,910 | 4.21 | 568,570 | 1,240 | 3.08 | 40,280 |
|  | No one speaks English as a main language | Female | 41,200 | 3.03 | 1,361,450 | 22,620 | 3.98 | 568,570 | 930 | 2.3 | 40,280 |
|  | Unknown | Female | 3,940 | 0.29 | 1,361,450 | 1,760 | 0.31 | 568,570 | 380 | 0.95 | 40,280 |
|  | All adults speak English as a main language | Male | 938,450 | 88 | 1,066,440 | 711,320 | 87.31 | 814,730 | 98,190 | 90.99 | 107,920 |
|  | At least one (but not all) adults speak/s English as a main language | Male | 52,510 | 4.92 | 1,066,440 | 41,570 | 5.1 | 814,730 | 4,220 | 3.91 | 107,920 |
|  | No adults speak English as a main language but at least one child does | Male | 35,360 | 3.32 | 1,066,440 | 31,620 | 3.88 | 814,730 | 2,690 | 2.49 | 107,920 |
|  | No one speaks English as a main language | Male | 37,140 | 3.48 | 1,066,440 | 27,900 | 3.42 | 814,730 | 1,540 | 1.43 | 107,920 |
|  | Unknown | Male | 2,970 | 0.28 | 1,066,440 | 2,320 | 0.28 | 814,730 | 1,280 | 1.18 | 107,920 |
| Household NS-SEC | Class 1.1 | Female | 56,150 | 4.12 | 1,361,450 | 12,220 | 2.15 | 568,570 | 1,010 | 2.51 | 40,280 |
|  | Class 1.2 | Female | 154,710 | 11.36 | 1,361,450 | 30,540 | 5.37 | 568,570 | 2,800 | 6.94 | 40,280 |
|  | Class 2 | Female | 344,900 | 25.33 | 1,361,450 | 96,710 | 17.01 | 568,570 | 6,800 | 16.88 | 40,280 |
|  | Class 3 | Female | 143,600 | 10.55 | 1,361,450 | 51,010 | 8.97 | 568,570 | 3,500 | 8.69 | 40,280 |
|  | Class 4 | Female | 192,570 | 14.14 | 1,361,450 | 78,560 | 13.82 | 568,570 | 5,410 | 13.42 | 40,280 |
|  | Class 5 | Female | 108,610 | 7.98 | 1,361,450 | 50,800 | 8.93 | 568,570 | 3,110 | 7.72 | 40,280 |
|  | Class 6 | Female | 158,600 | 11.65 | 1,361,450 | 94,420 | 16.61 | 568,570 | 6,370 | 15.82 | 40,280 |
|  | Class 7 | Female | 129,040 | 9.48 | 1,361,450 | 86,320 | 15.18 | 568,570 | 6,170 | 15.31 | 40,280 |
|  | Class 8 | Female | 48,920 | 3.59 | 1,361,450 | 53,540 | 9.42 | 568,570 | 3,970 | 9.84 | 40,280 |
|  | Students | Female | 20,170 | 1.48 | 1,361,450 | 12,470 | 2.19 | 568,570 | 750 | 1.86 | 40,280 |
|  | Unknown | Female | 4,180 | 0.31 | 1,361,450 | 1,980 | 0.35 | 568,570 | 400 | 1 | 40,280 |
|  | Class 1.1 | Male | 46,940 | 4.4 | 1,066,440 | 20,070 | 2.46 | 814,730 | 2,490 | 2.31 | 107,920 |
|  | Class 1.2 | Male | 131,350 | 12.32 | 1,066,440 | 50,520 | 6.2 | 814,730 | 7,170 | 6.64 | 107,920 |
|  | Class 2 | Male | 279,560 | 26.21 | 1,066,440 | 151,390 | 18.58 | 814,730 | 18,900 | 17.51 | 107,920 |
|  | Class 3 | Male | 111,060 | 10.41 | 1,066,440 | 76,950 | 9.44 | 814,730 | 9,520 | 8.82 | 107,920 |
|  | Class 4 | Male | 151,370 | 14.19 | 1,066,440 | 116,720 | 14.33 | 814,730 | 13,870 | 12.85 | 107,920 |
|  | Class 5 | Male | 84,880 | 7.96 | 1,066,440 | 73,530 | 9.03 | 814,730 | 9,150 | 8.48 | 107,920 |
|  | Class 6 | Male | 115,910 | 10.87 | 1,066,440 | 128,530 | 15.78 | 814,730 | 17,200 | 15.94 | 107,920 |
|  | Class 7 | Male | 95,070 | 8.91 | 1,066,440 | 116,170 | 14.26 | 814,730 | 16,380 | 15.17 | 107,920 |
|  | Class 8 | Male | 32,190 | 3.02 | 1,066,440 | 62,830 | 7.71 | 814,730 | 9,850 | 9.13 | 107,920 |
|  | Students | Male | 14,970 | 1.4 | 1,066,440 | 15,490 | 1.9 | 814,730 | 2,060 | 1.91 | 107,920 |
|  | Unknown | Male | 3,140 | 0.29 | 1,066,440 | 2,520 | 0.31 | 814,730 | 1,330 | 1.23 | 107,920 |
| Main language | Main language is English | Female | 1,284,370 | 94.34 | 1,361,450 | 532,790 | 93.71 | 568,570 | 37,550 | 93.21 | 40,280 |
|  | Main language is not English | Female | 53,120 | 3.9 | 1,361,450 | 29,730 | 5.23 | 568,570 | 1,890 | 4.7 | 40,280 |
|  | Unknown | Female | 23,960 | 1.76 | 1,361,450 | 6,050 | 1.06 | 568,570 | 840 | 2.09 | 40,280 |
|  | Main language is English | Male | 999,690 | 93.74 | 1,066,440 | 769,330 | 94.43 | 814,730 | 102,490 | 94.97 | 107,920 |
|  | Main language is not English | Male | 47,230 | 4.43 | 1,066,440 | 36,460 | 4.47 | 814,730 | 3,010 | 2.79 | 107,920 |
|  | Unknown | Male | 19,520 | 1.83 | 1,066,440 | 8,940 | 1.1 | 814,730 | 2,420 | 2.25 | 107,920 |
| Type of communal establishment | Communal establishment | Female | 3,810 | 0.28 | 1,361,450 | 1,720 | 0.3 | 568,570 | 370 | 0.92 | 40,280 |
|  | Household | Female | 1,357,640 | 99.72 | 1,361,450 | 566,850 | 99.7 | 568,570 | 39,910 | 99.08 | 40,280 |
|  | Communal establishment | Male | 2,850 | 0.27 | 1,066,440 | 2,250 | 0.28 | 814,730 | 1,260 | 1.17 | 107,920 |
|  | Household | Male | 1,063,590 | 99.73 | 1,066,440 | 812,470 | 99.72 | 814,730 | 106,660 | 98.83 | 107,920 |
| Unpaid carer status | Carer | Female | 42,380 | 3.11 | 1,361,450 | 25,220 | 4.44 | 568,570 | 1,190 | 2.94 | 40,280 |
|  | Non-carer | Female | 1,295,110 | 95.13 | 1,361,450 | 537,300 | 94.5 | 568,570 | 38,250 | 94.96 | 40,280 |
|  | Missing or not stated | Female | 23,960 | 1.76 | 1,361,450 | 6,050 | 1.06 | 568,570 | 840 | 2.09 | 40,280 |
|  | Carer | Male | 27,060 | 2.54 | 1,066,440 | 27,120 | 3.33 | 814,730 | 2,790 | 2.58 | 107,920 |
|  | Non-carer | Male | 1,019,850 | 95.63 | 1,066,440 | 778,660 | 95.57 | 814,730 | 102,710 | 95.17 | 107,920 |
|  | Missing or not stated | Male | 19,520 | 1.83 | 1,066,440 | 8,940 | 1.1 | 814,730 | 2,420 | 2.25 | 107,920 |

#### **Supplementary Table 5**: Secondary analysis: Characteristics of children by types of SEND among children with statements

| **Census characteristic** | **Level** | **Cognition and learning** | **Communication and interaction** | **Sensory and/or physical needs** | **Social, emotional and mental health** | **unresolved** |
| --- | --- | --- | --- | --- | --- | --- |
|  |  | **Count (%)** | **Count (%)** | **Count (%)** | **Count (%)** | **Count (%)** |
| ***Total*** | | ***58,120*** | ***42,040*** | ***14,870*** | ***23,690*** | ***9,490*** |
| Ethnic group (aggregated) | Asian/Asian British | 3620 (6.24) | 2290 (5.45) | 1630 (10.94) | 340 (1.45) | 430 (4.48) |
|  | Black/African/Caribbean/Black British | 2180 (3.75) | 2470 (5.88) | 580 (3.91) | 1050 (4.45) | 420 (4.37) |
|  | Mixed/multiple ethnic groups | 2150 (3.69) | 1850 (4.39) | 520 (3.52) | 1320 (5.55) | 400 (4.25) |
|  | Other ethnic group | 410 (0.71) | 300 (0.71) | 150 (0.99) | 80 (0.35) | 90 (0.91) |
|  | White | 48900 (84.14) | 34160 (81.26) | 11360 (76.38) | 20300 (85.69) | 7940 (83.71) |
|  | Unknown | 860 (1.48) | 970 (2.3) | 630 (4.25) | 590 (2.5) | 220 (2.28) |
| Country of birth | Non-UK | 2390 (4.1) | 1380 (3.27) | 930 (6.25) | 450 (1.88) | 450 (4.71) |
|  | UK | 54880 (94.42) | 39700 (94.43) | 13310 (89.49) | 22650 (95.61) | 8820 (93.01) |
|  | Missing or not stated | 860 (1.48) | 970 (2.3) | 630 (4.25) | 590 (2.5) | 220 (2.28) |
| Disability status | Day to day activities limited a lot/a little | 28380 (48.82) | 22870 (54.4) | 11630 (78.23) | 8400 (35.46) | 3820 (40.24) |
|  | Day to day activities not limited | 28890 (49.7) | 18200 (43.3) | 2610 (17.52) | 14700 (62.04) | 5450 (57.48) |
|  | Unknown | 860 (1.48) | 970 (2.3) | 630 (4.25) | 590 (2.5) | 220 (2.28) |
| Health status | Very bad health | 1940 (3.34) | 500 (1.18) | 920 (6.21) | 140 (0.61) | 160 (1.71) |
|  | Bad health | 3290 (5.66) | 1270 (3.02) | 1620 (10.92) | 650 (2.73) | 440 (4.65) |
|  | Fair health | 9690 (16.68) | 5290 (12.57) | 3740 (25.16) | 2880 (12.17) | 1410 (14.9) |
|  | Good health | 18100 (31.13) | 14000 (33.3) | 4560 (30.69) | 8150 (34.42) | 2820 (29.7) |
|  | Very good health | 24240 (41.71) | 20020 (47.62) | 3390 (22.77) | 11270 (47.56) | 4440 (46.77) |
|  | Unknown | 860 (1.48) | 970 (2.3) | 630 (4.25) | 590 (2.5) | 220 (2.28) |
| Household language | All adults speak English as a main language | 52480 (90.3) | 38050 (90.51) | 12630 (84.96) | 22090 (93.25) | 8640 (91.03) |
|  | At least one (but not all) adults speak/s English as a main language | 2580 (4.44) | 1860 (4.42) | 1020 (6.86) | 480 (2.04) | 320 (3.35) |
|  | No adults speak English as a main language but at least one child does | 1470 (2.53) | 1330 (3.15) | 610 (4.12) | 300 (1.25) | 230 (2.38) |
|  | No one speaks English as a main language | 1100 (1.89) | 530 (1.27) | 500 (3.37) | 130 (0.56) | 200 (2.13) |
|  | Unknown | 490 (0.84) | 280 (0.66) | 100 (0.7) | 690 (2.9) | 110 (1.11) |
| Household NS-SEC | Class 1.1 | 1160 (1.99) | 1280 (3.03) | 470 (3.17) | 360 (1.51) | 240 (2.56) |
|  | Class 1.2 | 3140 (5.41) | 4010 (9.54) | 1270 (8.53) | 880 (3.7) | 670 (7.04) |
|  | Class 2 | 8940 (15.37) | 8920 (21.23) | 3010 (20.24) | 3110 (13.14) | 1720 (18.09) |
|  | Class 3 | 4690 (8.08) | 4250 (10.12) | 1390 (9.37) | 1820 (7.67) | 870 (9.16) |
|  | Class 4 | 7930 (13.65) | 5210 (12.38) | 2090 (14.04) | 2880 (12.16) | 1170 (12.34) |
|  | Class 5 | 4990 (8.58) | 3490 (8.31) | 1210 (8.14) | 1830 (7.73) | 740 (7.83) |
|  | Class 6 | 9730 (16.74) | 6060 (14.42) | 2020 (13.61) | 4280 (18.05) | 1480 (15.64) |
|  | Class 7 | 10100 (17.37) | 5160 (12.27) | 1900 (12.76) | 3990 (16.86) | 1390 (14.7) |
|  | Class 8 | 5880 (10.11) | 2650 (6.31) | 1150 (7.75) | 3240 (13.69) | 890 (9.38) |
|  | Students | 1060 (1.83) | 720 (1.71) | 250 (1.65) | 590 (2.48) | 200 (2.1) |
|  | Unknown | 510 (0.88) | 290 (0.69) | 110 (0.73) | 710 (3.01) | 110 (1.15) |
| Main language | Main language is English | 55030 (94.67) | 40140 (95.48) | 13030 (87.63) | 22880 (96.6) | 8960 (94.45) |
|  | Main language is not English | 2240 (3.85) | 940 (2.22) | 1210 (8.12) | 210 (0.89) | 310 (3.27) |
|  | Unknown | 860 (1.48) | 970 (2.3) | 630 (4.25) | 590 (2.5) | 220 (2.28) |
| Type of communal establishment | Lives in a communal establishment | 470 (0.82) | 270 (0.63) | 100 (0.69) | 680 (2.89) | 100 (1.1) |
|  | Lives in a household | 57650 (99.18) | 41770 (99.37) | 14770 (99.31) | 23000 (97.11) | 9380 (98.9) |
| Unpaid carer status | Carer | 1670 (2.88) | 900 (2.15) | 340 (2.25) | 790 (3.32) | 280 (2.94) |
|  | Non-carer | 55590 (95.65) | 40170 (95.55) | 13900 (93.5) | 22310 (94.18) | 8990 (94.78) |
|  | Missing or not stated | 860 (1.48) | 970 (2.3) | 630 (4.25) | 590 (2.5) | 220 (2.28) |

#### **Supplementary table 6:** Secondary analysis: Characteristics of children with autism among children with Communication and Interaction SEND Type

| **Census characteristic** | **Level** | **Autism** | **Other subtypes of Communication and Interaction** |
| --- | --- | --- | --- |
|  |  | **Count (%)** | **Count (%)** |
| ***Total*** | | **25,270** | **16,770** |
| Ethnic group (aggregated) | Asian/Asian British | 1040 (4.10%) | 1260 (7.49%) |
|  | Black/African/Caribbean/Black British | 1380 (5.47%) | 1090 (6.51%) |
|  | Mixed/multiple ethnic groups | 1150 (4.57%) | 690 (4.12%) |
|  | Other ethnic group | 150 (0.60%) | 150 (0.88%) |
|  | White | 20880 (82.63%) | 13280 (79.20%) |
|  | Unknown | 660 (2.63%) | 300 (1.80%) |
| Country of birth | Non-UK | 770 (3.03%) | 610 (3.63%) |
|  | UK | 23840 (94.34%) | 15860 (94.57%) |
|  | Missing or not stated | 660 (2.63%) | 300 (1.80%) |
| Disability status | Day to day activities limited a lot/a little | 17320 (68.56%) | 5540 (33.06%) |
|  | Day to day activities not limited | 7280 (28.81%) | 10920 (65.14%) |
|  | Unknown | 660 (2.63%) | 300 (1.80%) |
| Health status | Very bad health | 400 (1.57%) | 100 (0.60%) |
|  | Bad health | 860 (3.40%) | 410 (2.46%) |
|  | Fair health | 3640 (14.41%) | 1640 (9.80%) |
|  | Good health | 8910 (35.24%) | 5090 (30.36%) |
|  | Very good health | 10800 (42.75%) | 9220 (54.97%) |
|  | Unknown | 660 (2.63%) | 300 (1.80%) |
| Household language | All adults speak English as a main language | 23260 (92.06%) | 14790 (88.17%) |
|  | At least one (but not all) adults speak/s English as a main language | 930 (3.68%) | 930 (5.53%) |
|  | No adults speak English as a main language but at least one child does | 650 (2.56%) | 680 (4.05%) |
|  | No one speaks English as a main language | 210 (0.82%) | 330 (1.94%) |
|  | Unknown | 230 (0.89%) | 50 (0.30%) |
| Household NS-SEC | Class 1.1 | 850 (3.36%) | 430 (2.55%) |
|  | Class 1.2 | 2750 (10.88%) | 1260 (7.52%) |
|  | Class 2 | 5900 (23.34%) | 3030 (18.05%) |
|  | Class 3 | 2730 (10.80%) | 1520 (9.09%) |
|  | Class 4 | 2940 (11.62%) | 2270 (13.53%) |
|  | Class 5 | 2050 (8.11%) | 1440 (8.60%) |
|  | Class 6 | 3420 (13.52%) | 2650 (15.77%) |
|  | Class 7 | 2710 (10.73%) | 2450 (14.59%) |
|  | Class 8 | 1260 (4.99%) | 1390 (8.29%) |
|  | Students | 440 (1.74%) | 280 (1.66%) |
|  | Unknown | 230 (0.91%) | 60 (0.35%) |
| Main language | Main language is English | 24150 (95.59%) | 15980 (95.31%) |
|  | Main language is not English | 450 (1.79%) | 480 (2.89%) |
|  | Unknown | 660 (2.63%) | 300 (1.80%) |
| Type of communal establishment | Lives in a communal establishment | 220 (0.86%) | 50 (0.29%) |
|  | Lives in a household | 25050 (99.14%) | 16720 (99.71%) |
| Unpaid carer status | Carer | 480 (1.88%) | 430 (2.55%) |
|  | Non-carer | 24130 (95.49%) | 16040 (95.65%) |
|  | Missing or not stated | 660 (2.63%) | 300 (1.80%) |

#### **Supplementary Table 7:** Main analysis. Three SEND levels: Crude and age-standardised rates per 100,000 person-years for death by suicide and count of suicides

| **Census characteristic** | **Level** | **No support** | | | **Support** | | | **Statement** | | |
| --- | --- | --- | --- | --- | --- | --- | --- | --- | --- | --- |
|  |  | **Count (n)** | **Crude rate per 100,000 person-years [95% CI]** | **Age-standardised rate per 100,000 person-years [95% CI]** | **Count (n)** | **Crude rate per 100,000 person-years [95% CI]** | **Age-standardised rate per 100,000 person-years [95% CI]** | **Count (n)** | **Crude rate per 100,000 person-years [95% CI]** | **Age-standardised rate per 100,000 person-years [95% CI]** |
| Sex | Female | 450 | 2.82 [2.56-3.08] | 2.82 [2.56-3.08] | 240 | 3.52 [3.07-3.97] | 3.51 [3.06-3.96] | 20 | 3.19 [1.79-5.26] | 3.18 [1.78-5.25] |
|  | Male | 850 | 6.74 [6.29-7.2] | 6.76 [6.31-7.22] | 960 | 9.98 [9.35-10.62] | 9.95 [9.32-10.58] | 110 | 8.95[7.3-10.6] | 8.77 [7.15-10.39] |
|  | Persons | 1300 | 4.55 [4.3-4.79] | 4.55 [4.31-4.8] | 1190 | 7.32 [6.91-7.74] | 7.29 [6.88-7.71] | 130 | 7.39 [6.11-8.67] | 7.25 [5.99-8.51] |
| Age on Census Day | 10 to 12 years | 390 | 4.26 [3.84-4.68] | [x] | 310 | 6.56 [5.83-7.29] | [x] | 20 | 4.7 [2.95-7.12] | [x] |
|  | 13 to 15 years | 480 | 4.98 [4.53-5.43] | [x] | 500 | 8.4 [7.66-9.14] | [x] | 60 | 8.97 [6.78-11.65] | [x] |
|  | 16 to 18 years | 430 | 4.39 [3.97-4.81] | [x] | 380 | 6.84 [6.15-7.52] | [x] | 50 | 7.81 [5.8-10.3] | [x] |
| Ethnic group (aggregated) | Asian/Asian British | 90 | 3.9 [3.14-4.79] | 3.89 [3.13-4.78] | 50 | 3.76 [2.76-5] | 3.79 [2.78-5.04] | [c] | [c] | [c] |
|  | Black/African/Caribbean/Black British | 50 | 4.44 [3.29-5.87] | 4.47 [3.3-5.91] | 40 | 3.93 [2.75-5.44] | 3.86 [2.7-5.35] | [c] | [c] | [c] |
|  | Mixed/multiple ethnic groups | 70 | 6.58 [5.17-8.26] | 6.71 [5.26-8.43] | 60 | 9.01 [6.93-11.53] | 8.95 [6.88-11.46] | 10 | 13.72 [6.58-25.22] | 14.02 [6.71-25.79] |
|  | Other ethnic group | [c] | [c] | [c] | [c] | [c] | [c] | [c] | [c] | [c] |
|  | White | 1060 | 4.57 [4.3-4.85] | 4.58 [4.3-4.86] | 1030 | 7.89 [7.41-8.37] | 7.85 [7.37-8.33] | 110 | 7.67 [6.24-9.1] | 7.49 [6.09-8.9] |
|  | Unknown | 20 | 2.93 [1.64-4.84] | 4 [1.71-7.37] | 10 | 6.25 [3.12-11.18] | 7.51 [3.59-13.7] | [c] | [c] | [c] |
| Country of birth | Non-UK | 110 | 5.46 [4.44-6.48] | 5.45 [4.44-6.47] | 80 | 7.01 [5.59-8.68] | 6.92 [5.51-8.58] | [c] | [c] | [c] |
|  | UK | 1170 | 4.51 [4.25-4.76] | 4.51 [4.26-4.77] | 1100 | 7.36 [6.93-7.8] | 7.33 [6.9-7.77] | 120 | 7.43 [6.1-8.75] | 7.28 [5.98-8.58] |
|  | Missing or not stated | 20 | 2.93 [1.64-4.84] | 4 [1.71-7.37] | 10 | 6.25 [3.12-11.18] | 7.51 [3.59-13.7] | [c] | [c] | [c] |
| Disability status | Day to day activities limited a lot/a little | 40 | 7.65 [5.44-10.45] | 7.52 [5.34-10.29] | 120 | 12.79 [10.46-15.12] | 12.77 [10.45-15.1] | 40 | 4.8 [3.46-6.49] | 4.82 [3.47-6.52] |
|  | Day to day activities not limited | 1240 | 4.52 [4.27-4.77] | 4.52 [4.27-4.78] | 1060 | 7.01 [6.59-7.43] | 6.98 [6.56-7.4] | 80 | 10.13 [8.07-12.56] | 9.7 [7.71-12.06] |
|  | Unknown | 20 | 2.93 [1.64-4.84] | 4 [1.71-7.37] | 10 | 6.25 [3.12-11.18] | 7.51 [3.59-13.7] | [c] | [c] | [c] |
| Health status | Very bad health | [c] | [c] | [c] | [c] |  |  | [c] | [c] | [c] |
|  | Bad health | [c] | [c] | [c] | 20 | 17.5 [10.37-27.66] | 17.44 [10.29-27.64] | [c] | [c] | [c] |
|  | Fair health | 40 | 10.16 [7.26-13.83] | 10.2 [7.21-13.98] | 60 | 11.09 [8.47-14.27] | 10.7 [8.16-13.78] | [c] | [c] | [c] |
|  | Good health | 240 | 4.71 [4.12-5.31] | 4.68 [4.09-5.28] | 290 | 7.41 [6.57-8.26] | 7.39 [6.54-8.24] | 50 | 8.78 [6.49-11.6] | 8.65 [6.39-11.45] |
|  | Very good health | 990 | 4.42 [4.15-4.7] | 4.42 [4.15-4.7] | 810 | 7.06 [6.57-7.54] | 7.01 [6.53-7.5] | 60 | 8.61 [6.63-10.99] | 8.37 [6.44-10.7] |
|  | Unknown | 20 | 2.93 [1.64-4.84] | 4 [1.71-7.37] | 10 | 6.25 [3.12-11.18] | 7.51 [3.59-13.7] | [c] | [c] | [c] |
| Household language | All adults speak English as a main language | 1160 | 4.6 [4.34-4.86] | 4.61 [4.34-4.87] | 1080 | 7.68 [7.22-8.14] | 7.65 [7.2-8.11] | 120 | 7.86 [6.47-9.25] | 7.72 [6.35-9.09] |
|  | At least one (but not all) adults speak/s English as a main language | 50 | 3.99 [2.99-5.2] | 3.97 [2.98-5.19] | 40 | 4.75 [3.4-6.47] | 4.66 [3.33-6.36] | [c] | [c] | [c] |
|  | No adults speak English as a main language but at least one child does | 40 | 3.82 [2.66-5.32] | 3.7 [2.55-5.17] | 30 | 4.6 [3.1-6.56] | 4.55 [3.04-6.54] | [c] | [c] | [c] |
|  | No one speaks English as a main language | 40 | 4.34 [3.1-5.92] | 4.47 [3.19-6.1] | 30 | 4.55 [3-6.62] | 4.56 [2.99-6.64] | [c] | [c] | [c] |
|  | Unknown | [c] | [c] | [c] | [c] | [c] | [c] | [c] | [c] | [c] |
| Household NS-SEC | Class 1.1 | 60 | 4.79 [3.63-6.19] | 4.79 [3.64-6.19] | 40 | 10.55 [7.54-14.36] | 10.47 [7.47-14.26] | [c] | [c] | [c] |
|  | Class 1.2 | 160 | 4.61 [3.88-5.34] | 4.63 [3.9-5.36] | 80 | 8.4 [6.66-10.46] | 8.38 [6.64-10.43] | 10 | 8.59 [4.12-15.79] | 8.67 [4.16-15.95] |
|  | Class 2 | 340 | 4.59 [4.1-5.08] | 4.59 [4.1-5.08] | 200 | 6.76 [5.82-7.7] | 6.72 [5.78-7.66] | 20 | 6.99 [4.33-10.69] | 7.09 [4.39-10.85] |
|  | Class 3 | 140 | 4.81 [4.03-5.6] | 4.81 [4.02-5.6] | 100 | 6.85 [5.53-8.18] | 6.87 [5.54-8.2] | 10 | 6.56 [3.15-12.06] | 6.51 [3.12-11.97] |
|  | Class 4 | 160 | 3.93 [3.32-4.54] | 3.93 [3.32-4.54] | 160 | 7.15 [6.06-8.25] | 7.12 [6.03-8.21] | 20 | 8.89 [5.43-13.73] | 8.75 [5.33-13.53] |
|  | Class 5 | 80 | 3.47 [2.75-4.33] | 3.49 [2.76-4.34] | 110 | 7.33 [5.94-8.72] | 7.32 [5.93-8.7] | 10 | 7.67 [3.83-13.72] | 7.21 [3.6-12.9] |
|  | Class 6 | 160 | 5.08 [4.31-5.86] | 5.1 [4.32-5.89] | 200 | 7.68 [6.61-8.74] | 7.64 [6.58-8.7] | 30 | 10.52 [7.04-15.06] | 10.04 [6.71-14.39] |
|  | Class 7 | 120 | 4.44 [3.64-5.25] | 4.46 [3.65-5.27] | 170 | 7.19 [6.11-8.27] | 7.15 [6.08-8.22] | 10 | 4.93 [2.63-8.44] | 4.85 [2.57-8.32] |
|  | Class 8 | 50 | 4.83 [3.53-6.44] | 4.94 [3.6-6.6] | 90 | 6.51 [5.23-8.02] | 6.53 [5.25-8.04] | [c] | [c] | [c] |
|  | Students | 30 | 7.75 [5.3-10.94] | 7.79 [5.32-11.01] | 30 | 8.22 [5.42-11.96] | 8.22 [5.41-11.96] | [c] | [c] | [c] |
|  | Unknown | [c] | [c] | [c] | 10 | 20.9 [10.43-37.4] | 36.08 [14.19-70.57] | [c] | [c] | [c] |
| Main language | Main language is English | 1230 | 4.57 [4.32-4.83] | 4.58 [4.32-4.83] | 1150 | 7.49 [7.05-7.92] | 7.46 [7.02-7.89] | 130 | 7.64 [6.3-8.98] | 7.49 [6.17-8.81] |
|  | Main language is not English | 60 | 4.66 [3.51-6.07] | 4.74 [3.57-6.18] | 30 | 4.37 [3.03-6.11] | 4.36 [3.01-6.09] | [c] | [c] | [c] |
|  | Unknown | 20 | 2.93 [1.64-4.84] | 4 [1.71-7.37] | 10 | 6.25 [3.12-11.18] | 7.51 [3.59-13.7] | [c] | [c] | [c] |
| Type of communal establishment | Communal establishment | [c] | [c] | [c] | [c] | [c] | [c] | [c] | [c] | [c] |
|  | Household | 1290 | 4.54 [4.29-4.78] | 4.54 [4.3-4.79] | 1180 | 7.29 [6.87-7.71] | 7.26 [6.85-7.67] | 130 | 7.35 [6.07-8.64] | 7.22 [5.96-8.48] |
| Unpaid carer status | Carer | 40 | 4.78 [3.4-6.53] | 5.35 [3.72-7.42] | 40 | 6.83 [4.92-9.24] | 6.9 [4.85-9.49] | [c] | [c] | [c] |
|  | Non-carer | 1240 | 4.57 [4.31-4.82] | 4.58 [4.32-4.83] | 1140 | 7.36 [6.93-7.78] | 7.33 [6.9-7.75] | 120 | 7.53 [6.2-8.85] | 7.39 [6.09-8.69] |
|  | Missing or not stated | 20 | 2.93 [1.64-4.84] | 4 [1.71-7.37] | 10 | 6.25 [3.12-11.18] | 7.51 [3.59-13.7] | [c] | [c] | [c] |

Note: Values [x] are not applicable for the given sample.
Values [c] have been suppressed for the purposes of statistical disclosure control.

#### **Supplementary table 8:** Secondary analysis. SEND types: Crude and age-standardised rates per 100,000 person-years for death by suicide and non-suicide deaths

| **Type of Special Educational Need and Disabilities** | **Suicide deaths** | | | **Non-suicide deaths** | | |
| --- | --- | --- | --- | --- | --- | --- |
|  | **Count of suicides** | **Crude rate per 100,000 person-years [95% CI]** | **Age-standardised rate per 100,000 person-years [95% CI]** | **Count of deaths by non-suicide death** | **Crude rate per 100,000 person-years [95% CI]** | **Age-standardised rate per 100,000 person-years [95% CI]** |
| Communication and interaction | 30 | 6.48 [4.24-8.73] | 6.63 [4.52-9.37] | 140 | 29.18 [24.42-33.95] | 29.18 [24.39-33.97] |
| Cognition and learning | 40 | 5.31 [3.58-7.05] | 5.18 [3.59-7.21] | 920 | 135.46 [126.69-144.22] | 144.09 [134.5-153.68] |
| Social, emotional and mental health | 40 | 15.5 [10.87-20.13] | 14.67 [10.57-19.81] | 170 | 59.48 [50.4-68.55] | 57.75 [48.8-66.71] |
| Sensory and/or physical needs | [c] | [c] | [c] | 410 | 236.01 [213.08-258.94] | 235.12 [212.26-257.99] |
| unresolved | 10 | 11.7 [5.34-18.05] | 12.84 [6.73-22.13] | 70 | 61.18 [46.64-75.72] | 62.71 [48.43-79.81] |

Values [c] Suppressed due to small counts (n < 10).

#### **Supplementary Table 9:** Secondary analysis. Autism Spectrum Disorder: Crude and age-standardised rates per 100,000 person-years for death by suicide

| **Autism Spectrum Disorder flag** | **Suicide deaths** | | | **Non-suicide deaths** | | |
| --- | --- | --- | --- | --- | --- | --- |
|  | **Count of suicides** | **Crude rate per 100,000 person-years [95% CI]** | **Age-standardised rate per 100,000 person-years [95% CI]** | **Count of deaths by non-suicide death** | **Crude rate per 100,000 person-years [95% CI]** | **Age-standardised rate per 100,000 person-years [95% CI]** |
| Autism Spectrum Disorder | 20 | 7.13 [4.28-9.98] | 7.32 [4.68-10.9] | 120 | 34.46 [28.19-40.73] | 34.72 [28.37-41.06] |
| No support | 1300 | 4.55 [4.3-4.79] | 4.55 [4.31-4.8] | 2970 | 10.39 [10.02-10.77] | 10.41 [10.04-10.79] |

#### **Supplementary Table 10:** Main analysis. Hazard ratios for deaths by suicide by Special Educational Needs and Disabilities category, with reference category “No support”

| **Special Educational Needs and Disabilities category** | **Model** | **Sex** | **Hazard ratio** | **Hazard ratio, 95% lower confidence limit** | **Hazard ratio, 95% upper confidence limit** | **p-value** |
| --- | --- | --- | --- | --- | --- | --- |
| Support | Age adjusted | Female | 1.23 | 1.05 | 1.44 | 0.01 |
| Statement | Age adjusted | Female | 1.15 | 0.69 | 1.92 | 0.59 |
| Support | Age adjusted | Male | 1.46 | 1.33 | 1.61 | <0.001 |
| Statement | Age adjusted | Male | 1.32 | 1.09 | 1.61 | 0.01 |
| Support | Age adjusted, NS-SEC | Female | 1.23 | 1.05 | 1.45 | 0.01 |
| Statement | Age adjusted, NS-SEC | Female | 1.14 | 0.68 | 1.91 | 0.62 |
| Support | Age adjusted, NS-SEC | Male | 1.46 | 1.33 | 1.60 | <0.001 |
| Statement | Age adjusted, NS-SEC | Male | 1.30 | 1.07 | 1.59 | 0.01 |
| Support | Age adjusted, health | Female | 1.17 | 0.99 | 1.37 | 0.06 |
| Statement | Age adjusted, health | Female | 0.87 | 0.51 | 1.48 | 0.60 |
| Support | Age adjusted, health | Male | 1.44 | 1.31 | 1.58 | <0.001 |
| Statement | Age adjusted, health | Male | 1.23 | 1.00 | 1.51 | 0.05 |
| Support | Age adjusted, disability status | Female | 1.19 | 1.02 | 1.40 | 0.03 |
| Statement | Age adjusted, disability status | Female | 0.78 | 0.45 | 1.36 | 0.38 |
| Support | Age adjusted, disability status | Male | 1.45 | 1.32 | 1.59 | <0.001 |
| Statement | Age adjusted, disability status | Male | 1.19 | 0.96 | 1.49 | 0.12 |
| Support | Age adjusted, main language | Female | 1.23 | 1.05 | 1.45 | 0.01 |
| Statement | Age adjusted, main language | Female | 1.15 | 0.69 | 1.93 | 0.58 |
| Support | Age adjusted, main language | Male | 1.46 | 1.33 | 1.60 | <0.001 |
| Statement | Age adjusted, main language | Male | 1.32 | 1.08 | 1.61 | 0.01 |
| Support | Age adjusted, communal establishment | Female | 1.23 | 1.05 | 1.44 | 0.01 |
| Statement | Age adjusted, communal establishment | Female | 1.14 | 0.68 | 1.91 | 0.61 |
| Support | Age adjusted, communal establishment | Male | 1.46 | 1.33 | 1.61 | <0.001 |
| Statement | Age adjusted, communal establishment | Male | 1.31 | 1.08 | 1.60 | 0.01 |
| Support | Age adjusted, unpaid carer status | Female | 1.23 | 1.05 | 1.44 | 0.01 |
| Statement | Age adjusted, unpaid carer status | Female | 1.15 | 0.69 | 1.93 | 0.58 |
| Support | Age adjusted, unpaid carer status | Male | 1.46 | 1.33 | 1.60 | <0.001 |
| Statement | Age adjusted, unpaid carer status | Male | 1.32 | 1.09 | 1.61 | 0.01 |
| Support | Age adjusted, ethnicity (aggregated) | Female | 1.23 | 1.05 | 1.44 | 0.01 |
| Statement | Age adjusted, ethnicity (aggregated) | Female | 1.16 | 0.69 | 1.94 | 0.57 |
| Support | Age adjusted, ethnicity (aggregated) | Male | 1.46 | 1.33 | 1.61 | <0.001 |
| Statement | Age adjusted, ethnicity (aggregated) | Male | 1.30 | 1.07 | 1.58 | 0.01 |
| Support | Age adjusted, ethnicity (aggregated), health | Female | 1.17 | 1.00 | 1.37 | 0.06 |
| Statement | Age adjusted, ethnicity (aggregated), health | Female | 0.87 | 0.51 | 1.48 | 0.61 |
| Support | Age adjusted, ethnicity (aggregated), health | Male | 1.44 | 1.31 | 1.58 | <0.001 |
| Statement | Age adjusted, ethnicity (aggregated), health | Male | 1.20 | 0.98 | 1.48 | 0.07 |
| Support | Age adjusted, ethnicity (aggregated), health, NS-SEC | Female | 1.17 | 0.99 | 1.38 | 0.06 |
| Statement | Age adjusted, ethnicity (aggregated), health, NS-SEC | Female | 0.87 | 0.51 | 1.48 | 0.60 |
| Support | Age adjusted, ethnicity (aggregated), health, NS-SEC | Male | 1.43 | 1.30 | 1.58 | <0.001 |
| Statement | Age adjusted, ethnicity (aggregated), health, NS-SEC | Male | 1.19 | 0.97 | 1.46 | 0.10 |

#### **Supplementary table 11:** Sensitivity analysis- restricted exposure to academic years 2008/2009 to 2010/2011. Hazard ratios for deaths by suicide by Special Educational Needs and Disabilities category, with reference category “No record of SEN”.

| **Special Educational Needs and Disabilities category** | **Model** | **Sex** | **Hazard ratio** | **Hazard ratio, 95% lower confidence limit** | **Hazard ratio, 95% upper confidence limit** | **p-value** |
| --- | --- | --- | --- | --- | --- | --- |
| Support | Age adjusted | Female | 1.40 | 1.18 | 1.66 | <0.001 |
| Statement | Age adjusted | Female | 1.12 | 0.65 | 1.95 | 0.68 |
| Support | Age adjusted | Male | 1.40 | 1.27 | 1.55 | <0.001 |
| Statement | Age adjusted | Male | 1.20 | 0.98 | 1.48 | 0.08 |

#### **Supplementary Table 12:** Sensitivity analysis - restricted exposure to academic years 2005/06 to 2010/11 and age 16 to 18 on Census day. Hazard ratios for deaths by suicide by Special Educational Needs and Disabilities category, with reference category “No support”

| **Special Educational Needs and Disabilities category** | **Model** | **Sex** | **Hazard ratio** | **Hazard ratio, 95% lower confidence limit** | **Hazard ratio, 95% upper confidence limit** | **p-value** |
| --- | --- | --- | --- | --- | --- | --- |
| Support & Statement | Age adjusted | Female | 1.48 | 1.09 | 2.01 | 0.01 |
| Support & Statement | Age adjusted | Male | 1.44 | 1.23 | 2.01 | <0.001 |

#### **Supplementary Table 13:** Sensitivity analysis. Modal SEND types: Crude and age-standardised rates per 100,000 person-years for death by suicide

| **Type of Special Educational Need and Disabilities** | **Suicide deaths** | | |
| --- | --- | --- | --- |
|  | **Count of suicides** | **Crude rate per 100,000 person-years [95% CI]** | **Age-standardised rate per 100,000 person-years [95% CI]** |
| Communication and interaction | 40 | 7.09 [4.94-9.86] | 7.31 [5.08-10.19] |
| Cognition and learning | 30 | 5.08 [3.52-7.1] | 5.01 [3.43-7.05] |
| Social, emotional and mental health | 40 | 14.98 [10.8-20.25] | 14.16 [10.16-19.18] |
| Sensory and/or physical needs | [c] | [c] | [c] |
| unresolved | 10.0 | 10.88 [5.62-19.01] | 12.05 [6.11-21.24] |

### 2. Supplementary text

#### **Supplementary text 1**: Derivation of SEND provision and SEND type categories

A) Derivation of SEND provision

SEN provision was derived from pupil-level school census records contained within the Growing Up in England (GUiE) Wave 3 datasets. These records originate from the Department for Education's Pupil Level Annual School Census (PLASC) and English School Census (ESC), which collect pupil characteristics and SEN provision information from schools and are incorporated into the National Pupil Database.

SEN status was recorded using different variables across academic years. In 2001/02, the variable senstage classified pupils according to the stage of SEN support received, ranging from no recorded SEN (code 0), through escalating levels of SEN “support” without a statement (codes 1–4), to a “statement” of SEN (code 5). In 2002/03–2004/05, the variable senstatus recorded whether pupils had no special provision (N), School Action (A), School Action Plus (P), School Action Plus with statutory assessment (Q), or a statement of SEN (S). From 2005/06 onwards, the variable senprovision recorded no identified SEN (N), School Action or Early Years Action (A), School Action Plus or Early Years Action Plus (P), and statemented SEN (S). To ensure comparability across years, these source variables were harmonised into three common categories: “no support”, “support”, and “statement”. The category "support" combined the staged support categories in senstage (codes 1–4), School Action, School Action Plus, and equivalent Early Years Action categories, while the category "statement" corresponded to pupils recorded as having a statement of SEN.

B) Derivation of SEND type categories

To derive the SEND types for children with a recorded SEND statement in our secondary analysis, we have examined records from academic years 2001/02 to 2010/11. We identified the most recently recorded SEND type and classified each case into one of five categories: (1) Cognition and learning difficulties, (2) Social, emotional and mental health needs, (3) Communication and interaction needs, (4) Physical or sensory needs, and (5) Unresolved - where multiple SEND types were recorded in the most recent year, or where the SEND type was missing or listed as 'Other'.

For the sensitivity analysis, we identified the most frequently occurring SEND type across the same time period and applied the same five-category classification. “Unresolved” - where more than one distinct SEND type occurred with equal frequency across the recorded years, or where the SEND type was missing or listed as 'Other'.

#### **Supplementary text 2**: Sensitivity analysis – restricted exposure to academic years 2008/2009 to 2010/2011. Hazard ratios for deaths by suicide by SEND status.

Restricting the exposure period for identifying SEND provision to academic years 2008/09 to 2010/11 resulted in modest changes to the estimated associations (Supplementary Table 11). Among males, the hazard ratio for those receiving SEND support decreased slightly from 1.46 in the main analysis to 1.40, while the hazard ratio for those with SEND statements decreased from 1.32 to 1.20 and was no longer statistically significant (p = 0.08). Among females, the association for those receiving SEND support was stronger than in the main analysis (HR 1.40 versus 1.23), whereas the estimate for those with SEND statements remained largely unchanged (HR 1.12 versus 1.15) and was not statistically significant.

#### **Supplementary text 3**: Sensitivity analysis – restricted exposure to academic years 2005/06 to 2010/11 and age 16 to 18 on Census day. Hazard ratios for deaths by suicide by SEND status.

Restricting the sample to individuals aged 16 to 18 years on Census Day and limiting the exposure period to academic years 2005/06 to 2010/11 reduced the sample size and required combining SEND support and statements into a single category because of small counts. Compared with individuals with no SEND provision, those with a record of SEND support or a statement had an increased risk of death by suicide among both males (HR 1.44, 95% CI 1.23 to 2.01) and females (HR 1.48, 95% CI 1.09 to 2.01) (Supplementary Table 12). These estimates were similar in magnitude to those observed in the main analysis for SEND support, indicating that the overall findings were robust to restricting the study population and exposure period.

#### **Supplementary text 4**: Sensitivity analysis - calculation of crude and age-standardised rates based on the most frequently recorded type of SEND.

The ranking of suicide rates by SEND type remained consistent with the main analysis (Supplementary Table 13). Individuals with social, emotional and mental health needs had the highest crude suicide rate (14.98 per 100,000 person-years), followed by those with communication and interaction needs (7.09 per 100,000 person-years) and cognition and learning difficulties (5.08 per 100,000 person-years). The unresolved category also showed an elevated crude suicide rate (10.88 per 100,000 person-years). Counts in the sensory and/or physical needs category were too small for disclosure. Compared with the main analysis, only minor changes in the estimated rates were observed, with a slight decrease for social, emotional and mental health needs (15.50 to 14.98 per 100,000 person-years) and a modest increase for communication and interaction needs (6.48 to 7.09 per 100,000 person-years). Overall, the pattern and ranking of suicide rates across SEND types remained unchanged.

### 3. Supplementary Figure

#### **Supplementary Figure 1:** Population and linkage flow diagram.

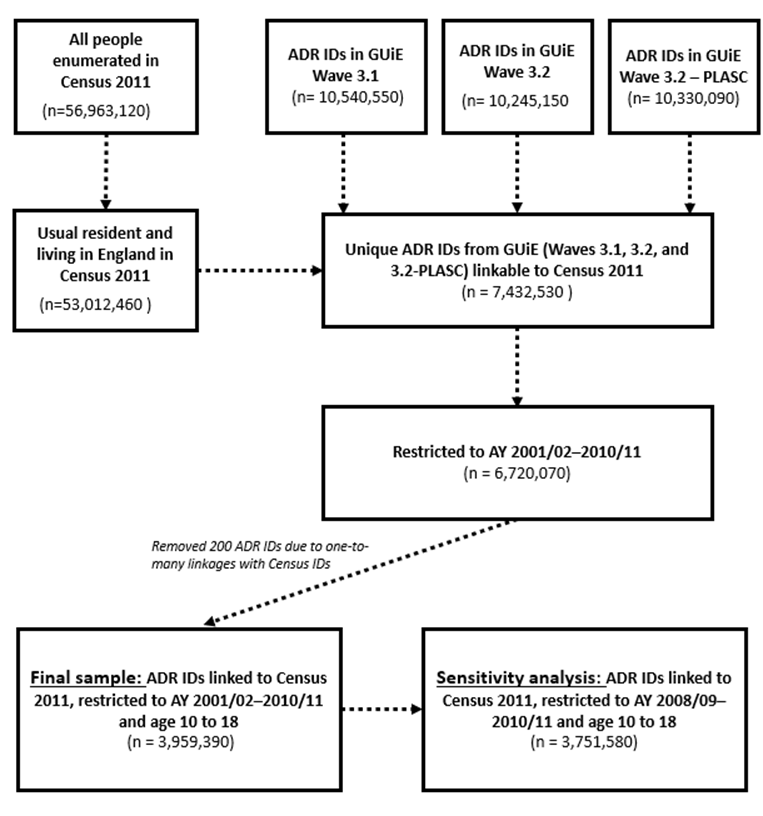

*Note: GUIE waves (3.1, 3.2, 3.2- Pupil level annual school census (PLASC)) represent sequential waves of linked administrative data. Each wave links education records to cohorts of children and extends the dataset further back in historical academic years. GUIE wave 3.1 includes data from academic years 2010/2011 to 2014/2015, wave 3.2 includes data from academic years 2005/2006 to 2009/2010, and wave 3.2-PLASC* includes data from academic years 2001/2002 to 2005/2006.
